# Disruption in the Host-Phage Dynamics and Altered Microbial Diversity in the Upper Respiratory Tract of SARS-CoV-2 Infected Individuals

**DOI:** 10.1101/2025.03.25.25324602

**Authors:** Siddharth Singh Tomar, Krishna Khairnar

**Author notes:** Corresponding author: Correspondence to Krishna Khairnar.

## Abstract

The Upper Respiratory Tract (URT) is an important site for the predisposition and multiplication of the SARS-CoV-2 virus. Therefore, URT is a critical site to investigate the changes in the microbiome caused by the SARS-CoV-2 infection. In this study, we have used the whole genome shotgun metagenomic approach to investigate URT swab samples (*n=96*) collected from SARS-CoV-2-positive individuals (*n=48*) (non-hospitalised but symptomatic) and healthy controls (*n=48*) belonging to five districts of central India. This study aims to compare the phageome diversity and investigate the correlation of the phageome profiles with the sample type (SARS-CoV-2 or Control) to determine the nature of phage-host interactions and to assess the effect of SARS-CoV-2 viral load over host and phage abundance. The results showed that *Detrevirus* was a prominent bacteriophage in controls and *Maxrubnervirus* in SARS-CoV-2 samples. Higher Chao1 indices were observed in the SARS-CoV-2 group for bacteria (886.00 vs. 351.00, p < 0.0001) and phages (39.00 vs. 16.00, p = 0.0002). The Simpson index was lower for bacteria (0.88 vs. 0.93, p = 0.0024) and higher for phages (0.86 vs. 0.79, p = 0.0384). Ct-dependent variations in bacterial (H = 6.69, p = 0.035) and phage (H = 8.97, p = 0.011) abundances were also observed. Disrupted host-phage interactions were observed in SARS-CoV-2 samples, with a weaker model fit (logistic R^2^ = 0.7425) than controls (logistic R^2^ = 0.9265). These findings highlight the need to integrate virome and bacteriome analyses with potential diagnostic, prognostic, and therapeutic applications for infectious disease research.

## 1. Introduction

The COVID-19 pandemic caused by the SARS-CoV-2 virus has caused a severe loss of life and productivity around the globe. The SARS-CoV-2 virus is transmitted mainly through breathing or direct contact with virus-containing droplets and aerosols. The virus enters the host’s body through the upper respiratory tract (URT) and shows its initial predisposition, infecting the epithelial cells through hACE2 receptors. After multiplying in the URT, the virus moves towards the lower respiratory tract and the lungs, causing severe symptoms. The role of URT in the predisposition and multiplication of the virus makes it a critical site to investigate the changes in the microbiome caused by the SARS-CoV-2 infection.

The effect of respiratory viral infections on URT and the gut microbiome is widely studied. A growing body of evidence suggests that the SARS-CoV-2 infection could increase the overall microbiome diversity compared to healthy controls [1]. Specific bacterial taxa, such as *Peptoniphilus lacrimalis* and *Campylobacter hominis*, are more abundant in infected individuals [2]. A recent study also reported SARS-CoV-2 infection-induced dysbiosis of the URT microbiome and the changes in the URT microbiome corresponding to the severity of the infection [3]. Studies have shown a correlation between the SARS-CoV-2 viral load and the diversity of the URT microbiome and the microbiome profiles, showing variant-specific patterns along different waves of COVID-19 pandemics [4,5]. Age-dependent variations in the microbiome diversity and abundance were also reported among SARS-CoV-2 infected individuals, showing the role of the respiratory microbiome in modulating the host response and susceptibility against the disease [6].

The effect of SARS-CoV-2 infection on the respiratory microbiome is quite evident in the growing body of knowledge in this domain. However, very few studies are available on a crucial subset of the URT microbiome, i.e., the phageome. The part of the microbiome that collectively represents the taxa belonging to bacteriophages is known as the phageome.

The changes in the URT phageome corresponding to SARS-CoV-2 infection were not discussed directly in most studies related to the URT microbiome. A metagenomic study by Lu et al. (2021) has investigated the Compositional changes in the Intestinal DNA Virome of SARS-CoV-2 infected individuals; the authors have observed significant changes in the intestinal DNA Virome but could not draw any significant correlation between disease progression and the alterations in the DNA virome [7]. Another study with a similar objective to assess the impact of alteration in gut virome was done by Zuo et al. (2021), in which the authors observed the differential abundance and composition of DNA and RNA virome among SARS-CoV-2 and control groups; the authors have also observed the potential correlation between disease severity and the changes in the gut virome [8].

Aspects such as the therapeutic potentials of bacteriophages in the management of SARS-CoV-2 infection and using phage-displayed receptor binding domains for modelling interactions with SARS-CoV-2 spike proteins were explored in certain studies [9,10]. Ferravante et al. (2022) have attempted to elucidate the URT phageome diversity among different waves of the COVID-19 pandemic using the RNA-seq metagenomics approach. The findings suggest that the severity of COVID-19 infection correlated with the abundance of Caudovirales in the URT swab samples. This study is one of the few studies that have attempted to use the metagenomic approach to assess the URT phageome of SARS-CoV-2-infected individuals [11].

Metagenomics has significantly enhanced the ability to understand the viromes in clinical and epidemiological settings. Metagenomics facilitates the exhaustive profiling of viromes without prior compositional knowledge, which helps identify novel and low-abundant viral sequences. Metagenomics also overcome the limitations of culture-dependent methods, particularly in viromics [12,13]. These advantages of the metagenomics approach make it a method of choice for surveillance and public health applications.

In this study, we have used the whole genome shotgun metagenomic approach to investigate URT swab samples (*n=96*) collected from SARS-CoV-2-positive individuals (non-hospitalised but symptomatic) and healthy controls belonging to five districts of central India. This study aims to compare the phageome diversity and investigate the correlation of the phageome profiles with the sample type (SARS-CoV-2 or Control) to determine the nature of phage-host interactions and to assess the effect of SARS-CoV-2 viral load over host and phage abundance among the infected samples.

## 2. Materials and Methods

URT swab samples (n=96) were collected in Viral Transport Medium (VTM) during March-April 2023 from the five districts of central India, Namely Nagpur, Wardha, Gadchiroli, Chandrapur, and Bhandara. The samples were divided into two groups, i.e., the SARS-CoV-2 Positive Group and The SARS-CoV-2 Negative Group (Control), having a median age of 36 years (IQR: 19–67) and 29 years (IQR: 22–37), respectively. The SARS-CoV-2 group belongs to symptomatic but non-hospitalized RTPCR-positive (Ct <25) participants with severe acute respiratory infection (SARI) or influenza-like illness (ILI) symptoms. Control participants were asymptomatic and RTPCR-negative. The collected samples were stored at 4°C for ≤5 days (short-term) or -80°C (long-term). Processing of collected samples for RTPCR testing was done under Biosafety level-II conditions.

DNA extraction for metagenomic sequencing was carried out by using the QIAamp DNA Microbiome Kit. The quality check for DNA concentration was done using a Qubit fluorometer, and purity analysis was done using a Nanodrop spectrophotometer by examining A260/280 and A260/230 ratios. A QIAseq FX DNA Library Preparation Kit was used to prepare the metagenomic library, and the Illumina NextSeq550 platform with (2×150 bp high-output kit) generating paired-end reads over 300 cycles was used for metagenomic next-generation sequencing.

Metagenomic data analysis was done using the Chan Zuckerberg ID (CZID) web-based Illumina mNGS Pipeline v8.3 [14]. Preliminary Sequencing data quality control (QC) was done using MultiQC, and further, QC involved removing Sequencing artefacts such as adapters, low-quality sequences, and low-complexity reads using the fastp tool. External RNA Controls Consortium (ERCC) sequences were removed using the Bowtie2 tool. Human reads were also removed using Bowtie2 and HISAT2, and the remaining non-host reads were aligned to the NCBI nucleotide and protein databases (NCBI Index Date: 2024-02-06) using Minimap2 and Diamond tools, respectively.

SPAdes was used for de novo genome assembly, and the assembled contigs were then further aligned using Bowtie2. The BLAST tool was used for annotation and taxon count generation. The statistical analysis and visualization of the abundance and compositional data were performed using Python 3.10.12 and the following packages: SciPy (1.13.1) for t-tests and ANOVA. statsmodels (0.14.4) for regression models and hypothesis testing. scikit-learn (1.5.2) for PCA, PCoA, and multiple regression. matplotlib (3.7.1) & seaborn (0.13.2) were used for heatmaps and violin plots. Plotly (5.24.1) for interactive visualizations. Pandas (2.2.2) was used for data transformation, and numpy (1.26.4) for statistical analysis. The Reads per million (RPM) was used as the abundance metric for analysis and visualisation. The virus-host association data was accessed using the Virus-Host DB database (Index Date: November 23, 2024) [15]

## 3. Results

### 3.1. Compositional differences between SARS-CoV-2 and control groups

The comparative visualization **(Figure 1)** represents bacterial and phage genera distributions in control and SARS-CoV-2 samples. The top bacterial genus in both groups is *Streptococcus*, while *Sphingomonas, Orchrobactrum*, and *Brevundimonas* appear in SARS-CoV-2 samples but not in controls. The top phage genus in the control group is *Detrevirus*, whereas *Maxrubnervirus* is the most abundant in SARS-CoV-2 samples. After applying the abundance threshold of > 10 RPM for bacteria and > 1 RPM for bacteriophage, 34 bacterial genera were found in controls and 88 in SARS-CoV-2 samples, with 24 shared genera between both groups. For phages, 28 genera are found in controls and 79 in SARS-CoV-2 samples, with 16 genera common to both groups **(Figure 1)**. The comparative heatmaps **(Figure 2)** illustrate the log-transformed abundance of bacteria and bacteriophages on the genus level in the control and SARS-CoV-2 groups across all the samples.

**Figure 1:**
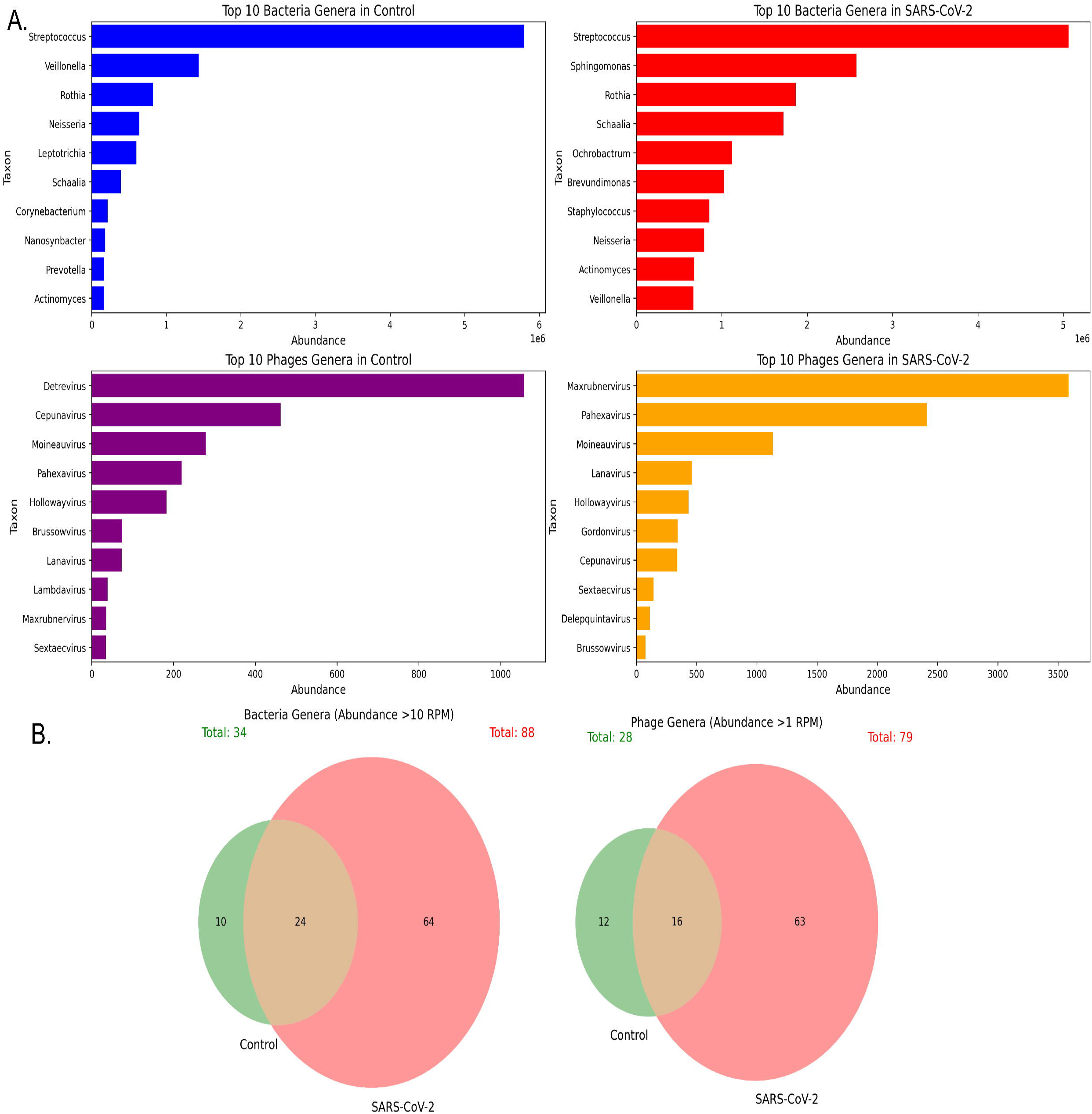
Variations in the upper respiratory tract bacterial and phage diversity in SARS-CoV-2-infected individuals and healthy controls. (A) Bar plots representing the top 10 most abundant bacterial (top row) and phage (bottom row) genera in control (left) and SARS-CoV-2-infected (right) samples. (B) Venn diagrams showing the number of bacterial (left) and phage (right) genera detected in control and SARS-CoV-2-infected samples.

**Figure 2:**
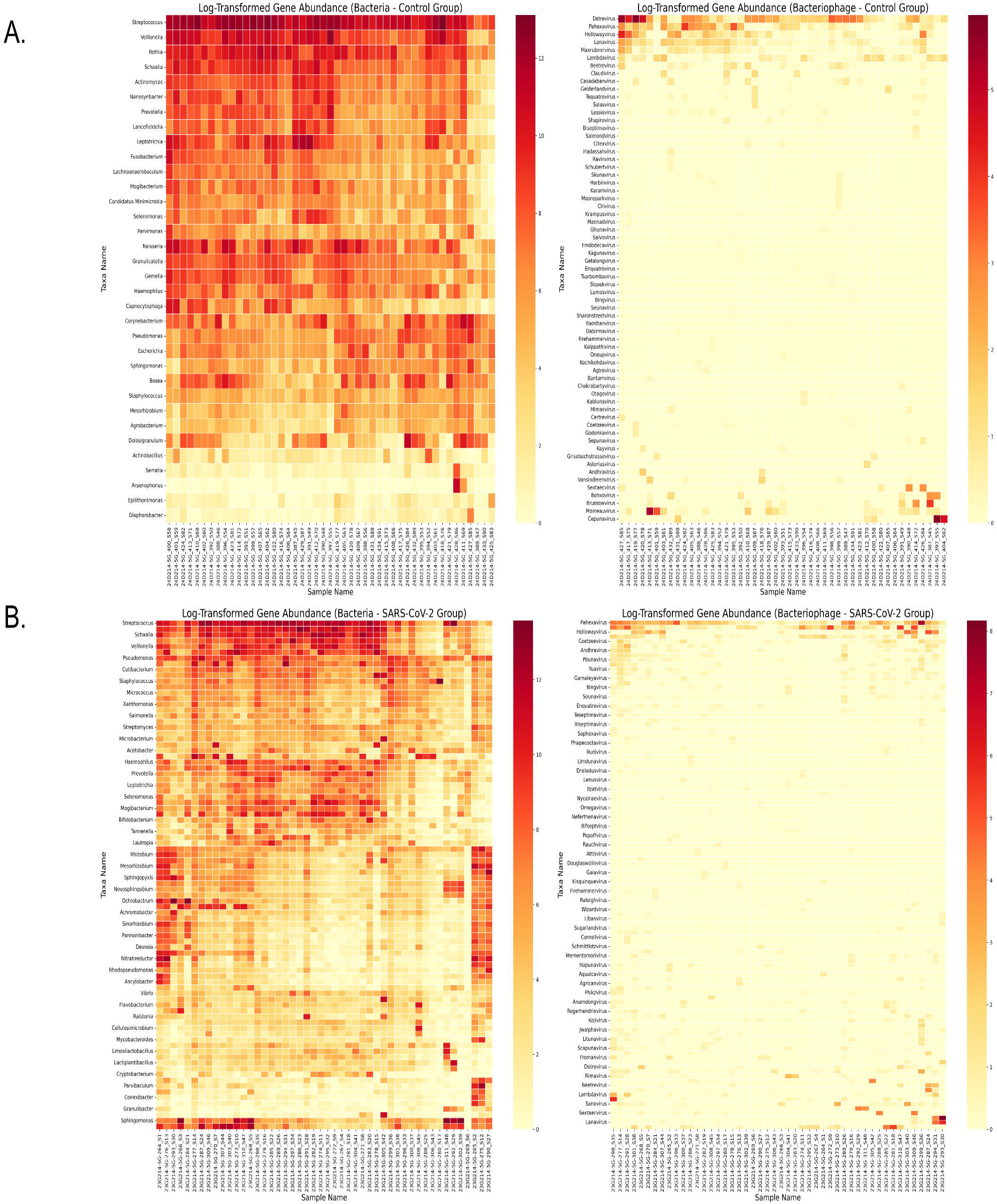
Heatmaps of log-transformed abundance in the control and SARS-CoV-2 groups. (A) Heatmaps depicting the log-transformed abundance of bacterial (left) and bacteriophage (right) taxa in the control group. Each row represents a taxon, and each column corresponds to a sample. (B) Heatmaps showing the log-transformed abundance of bacterial (left) and bacteriophage (right) taxa in the SARS-CoV-2 group.

A comparative host-phage association analysis was also performed on the species level data **(Figure 3)**, showing that host bacterial abundance in the control group follows a structured distribution, with some dominant taxa such as *Streptococcus thermophilus* and *Pseudomonas aeruginosa*. The phage abundance in the control group remains less pronounced and with limited diversity. Meanwhile, the SARS-CoV-2 group exhibits a more consistent and pronounced bacterial abundance. The abundance of Host bacterial species in the SARS-CoV-2 group appears to be more dispersed across samples, and phage abundance shows a broader distribution with more taxa detected than the control group.

**Figure 3:**
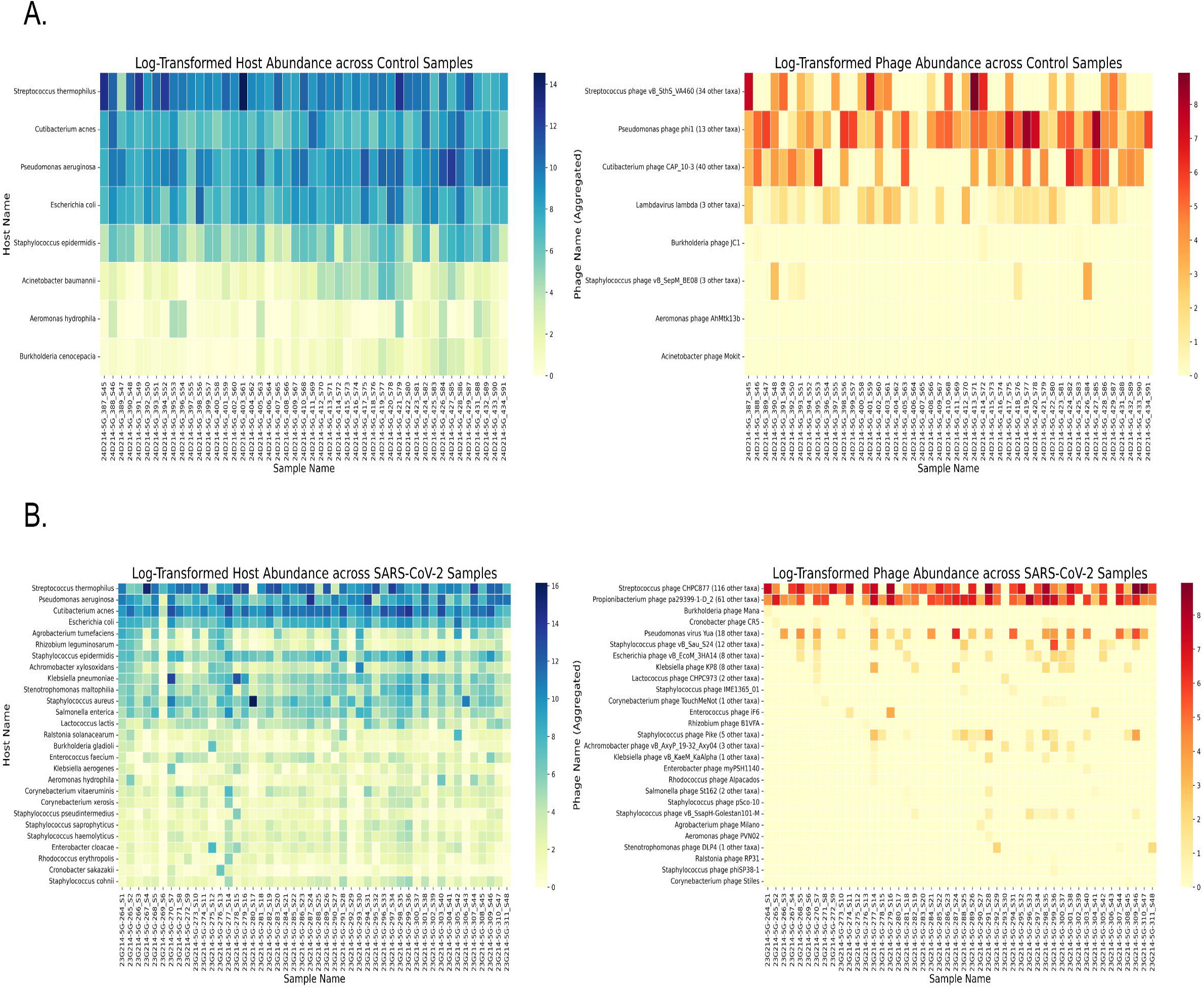
Heatmaps of log-transformed host and phage abundance in the control and SARS-CoV-2 groups. (A) Heatmaps representing log-transformed bacterial host abundance (left) and bacteriophage abundance (right) across control samples. Each row corresponds to a bacterial host or phage taxon, and each column represents a sample. (B) Heatmaps showing log-transformed bacterial host abundance (left) and phage abundance (right) across SARS-CoV-2 samples.

### 3.2. Alpha and Beta Diversity

The Chao1 index was significantly higher in the SARS-CoV-2 group (886.00) for bacterial diversity when compared to controls (351.00, *p* < 0.0001). At the same time, the Simpson index for bacterial diversity was significantly lower in the SARS-CoV-2 group (0.88) compared to controls (0.93, *p* = 0.0024). For the bacteriophage diversity, the chao1 index is higher in the SARS-CoV-2 group (39.00) compared to the control (16.00, p = 0.0002). The Simpson index was also higher in the SARS-CoV-2 group (0.86) than in controls (0.79, p = 0.0384). **(Figure 4)**

**Figure 4:**
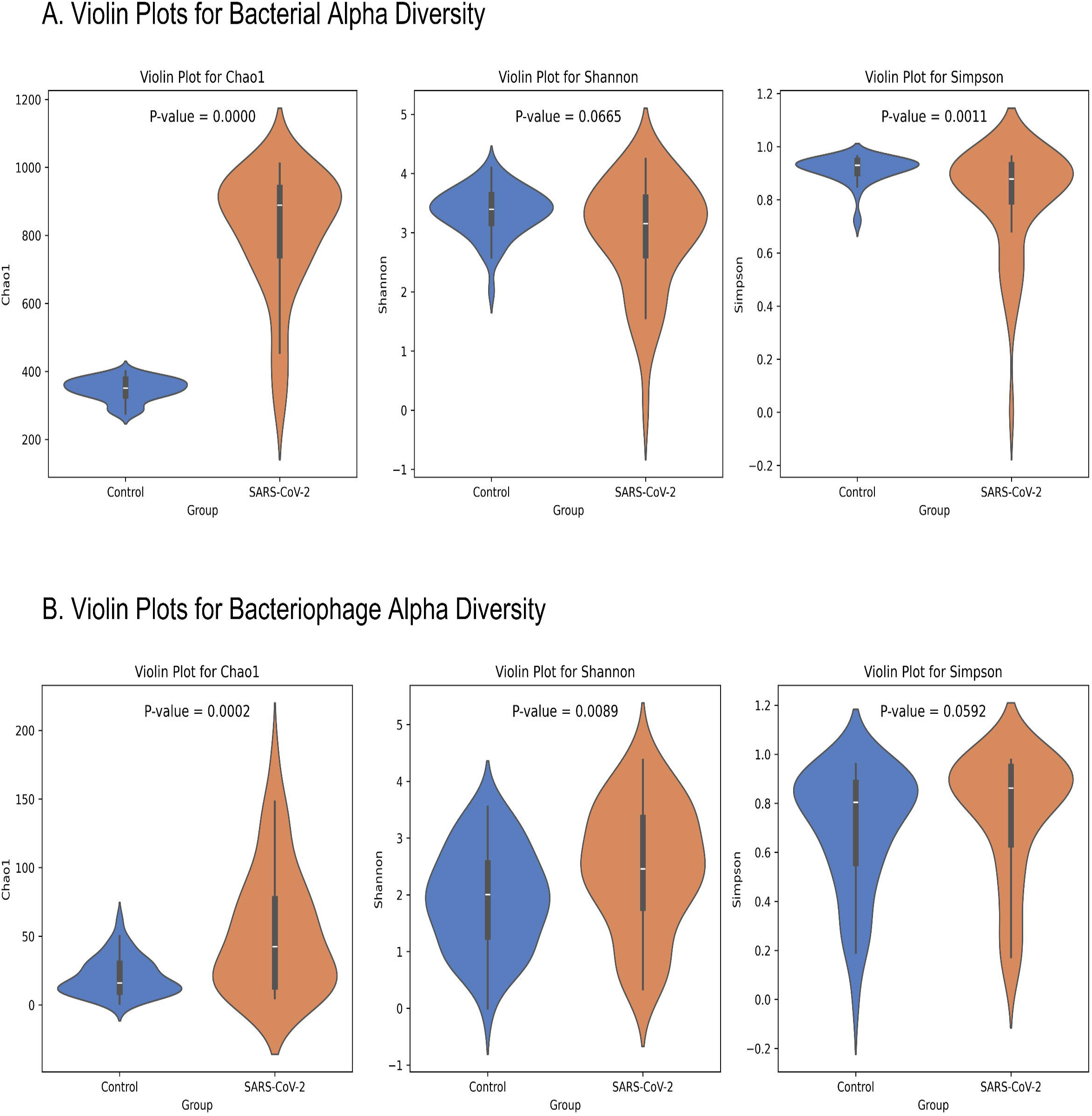
Violin plots comparing bacterial and bacteriophage alpha diversity between control and SARS-CoV-2 groups. (A) Violin plots depicting bacterial alpha diversity using Chao1, Shannon, and Simpson indices in control (blue) and SARS-CoV-2 (orange) groups. (B) Violin plots representing bacteriophage alpha diversity using the same three indices.

The top-left section of the Bray Curtis dissimilarity heatmap contains SARS-CoV-2 samples (red sample tags), and the bottom-right section consists of control samples (blue sample tags). **(Figure 5)** The dark blue diagonal line indicates 100% similarity (self-comparison), while the red shades indicate dissimilarities. Small blue regions within the red quadrants and vice versa indicate shared features among specific samples. The phage group (Panel A) with SARS-CoV-2 samples exhibited greater dispersion and no distinct clustering pattern. However, the bacterial community (Panel B) shows a prominent separation between the two groups.

**Figure 5:**
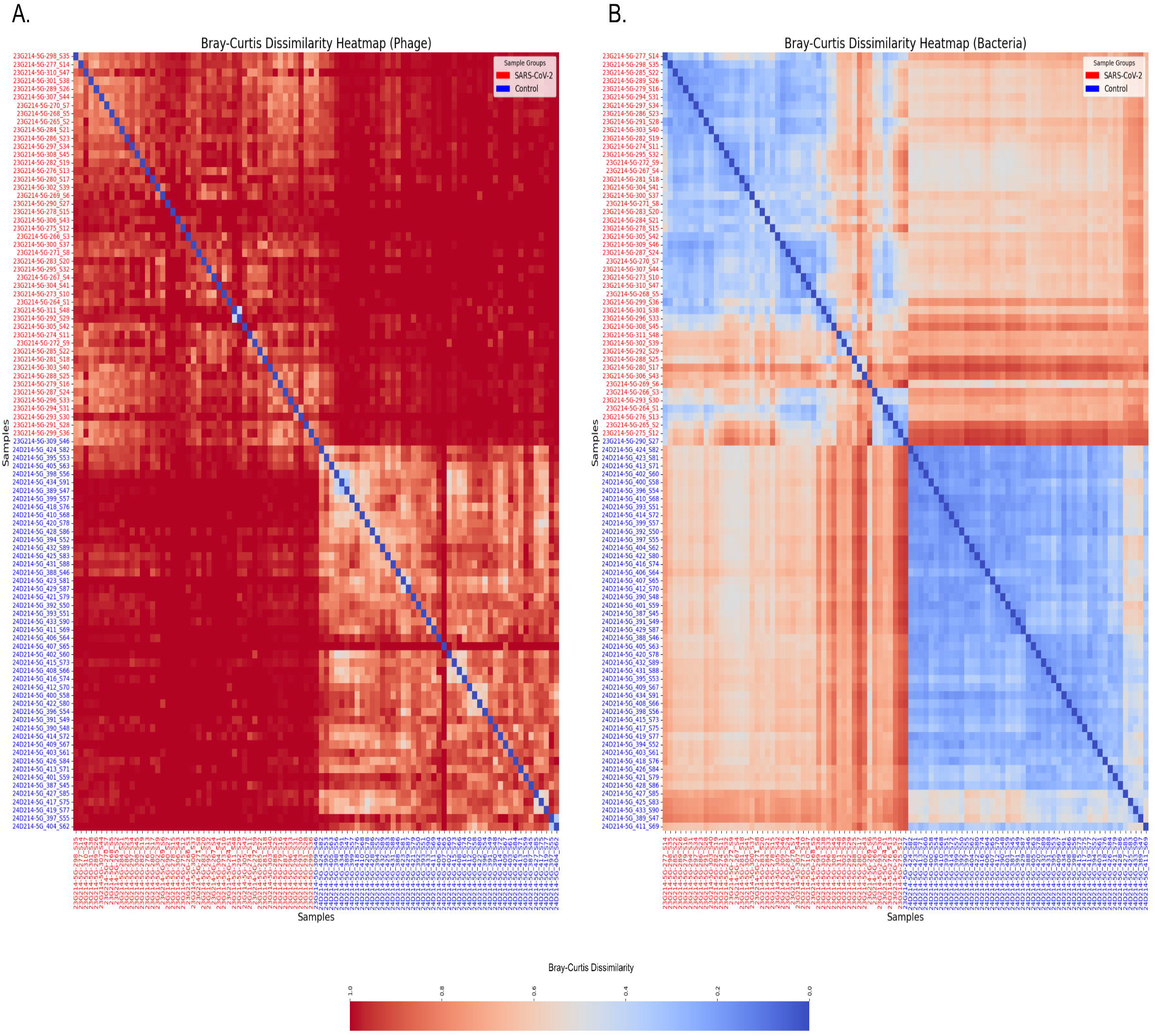
Bray-Curtis Dissimilarity Heatmap of Microbial Communities. Panel (A) Bray-Curtis dissimilarity heatmap represent the phage community composition across SARS-CoV-2 (red-labeled) and control (blue-labeled) samples. Similarly, Panel (B) Bray-Curtis dissimilarity heatmap represents the bacterial community composition among SARS-CoV-2 and Control samples.

The Principal Coordinates Analysis (PCoA) plots **(Figure 6)** also show differences in bacterial and bacteriophage community compositions between SARS-CoV-2 and control groups. In the Bacteria PCoA Plot (A), the PCoA Axis 1 (23.07%) and Axis 2 (13.51%) indicate a clear distinction between the two groups, with SARS-CoV-2 samples (red) clustering separately from control samples (blue). For Bacteriophage PCoA Plot (B), the separation along PCoA Axis 1 (9.10%) and Axis 2 (3.98%) is less prominent than in the bacterial plot, but a distinct clustering is still visible. The SARS-CoV-2 samples (red) appear more tightly clustered, while the control samples (blue) show more significant divergence.

**Figure 6:**
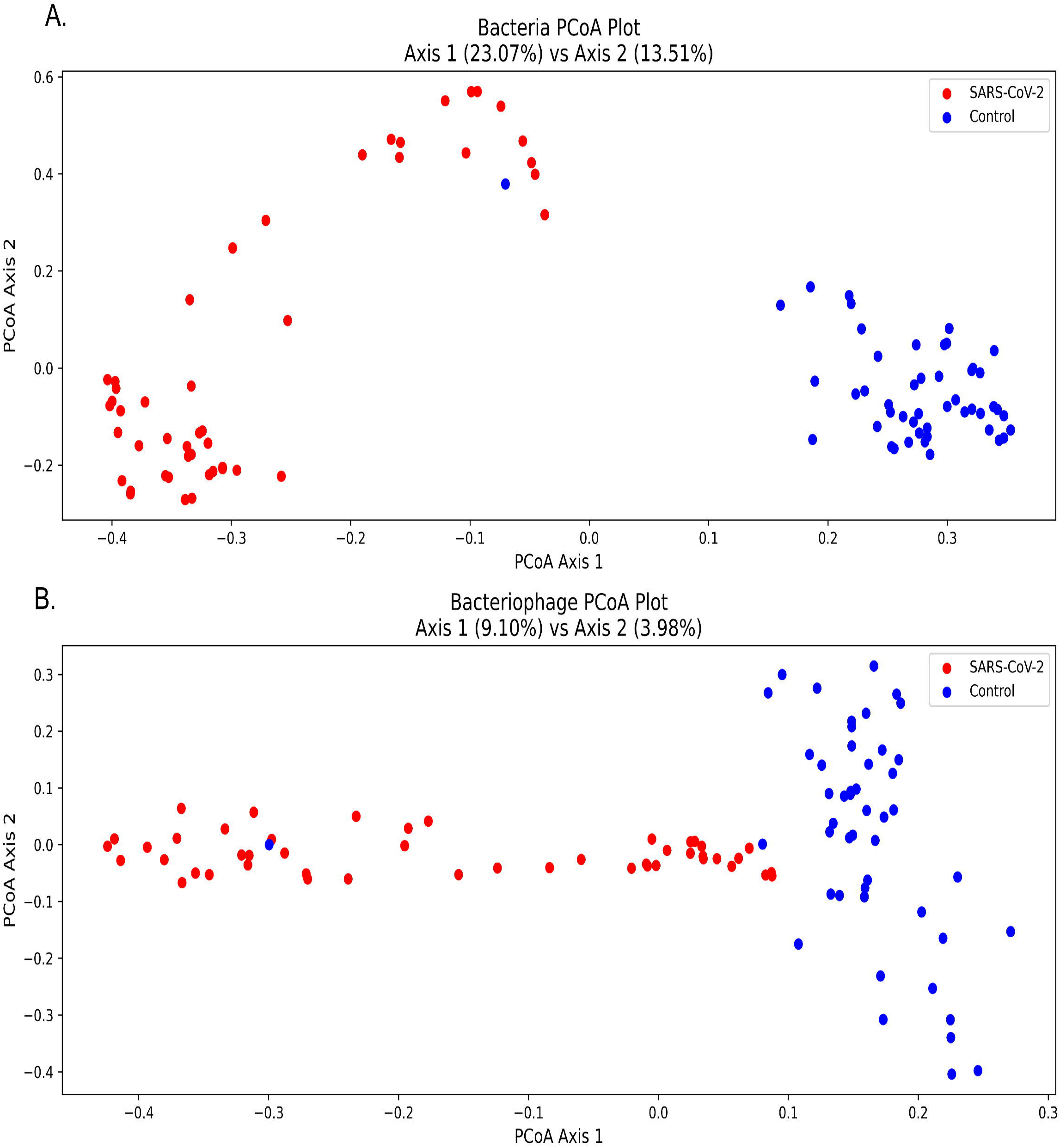
Principal Coordinates Analysis (PCoA) of Bacterial and Bacteriophage Communities. Panel (A) represents the PCoA plot of bacterial community composition based on Bray-Curtis dissimilarity. Each point represents a sample, with SARS-CoV-2 samples in red and control samples in blue. Panel (B) represents the PCoA plot of bacteriophage community composition using Bray-Curtis dissimilarity.

### 3.3. Effect of SARS-CoV-2 viral load (Ct value) on host and phage abundance

To assess the effect of the cycle threshold value (Ct value) on host bacterial abundance and corresponding mean phage abundance in the samples belonging to the SARS-CoV-2 group, we have used the Kruskal-Wallis test. The results showed statistically significant differences in Mean Phage Abundance (H = 8.97, *p* = 0.011) and Bacteria Abundance (H = 6.69, *p* = 0.035) across the three Ct value categories: Low Ct (High Viral Load, Ct < 20), Medium Ct (Moderate Viral Load, Ct 20–24), and High Ct (Low Viral Load, Ct > 24). Dunn’s post-hoc test for Bacteria Abundance indicated a significant difference between High and Medium Ct categories (*p* = 0.038), while comparisons between High vs. Low (*p* = 0.784) and Medium vs. Low (*p* = 0.591) were not significant. For Mean Phage Abundance, Dunn’s post-hoc test showed significant differences between High vs. Low (*p* = 0.021) and Medium vs. Low (*p* = 0.028), with no significant difference between High and Medium Ct categories (*p* = 1.000). **(Figure 7)**

**Figure 7:**
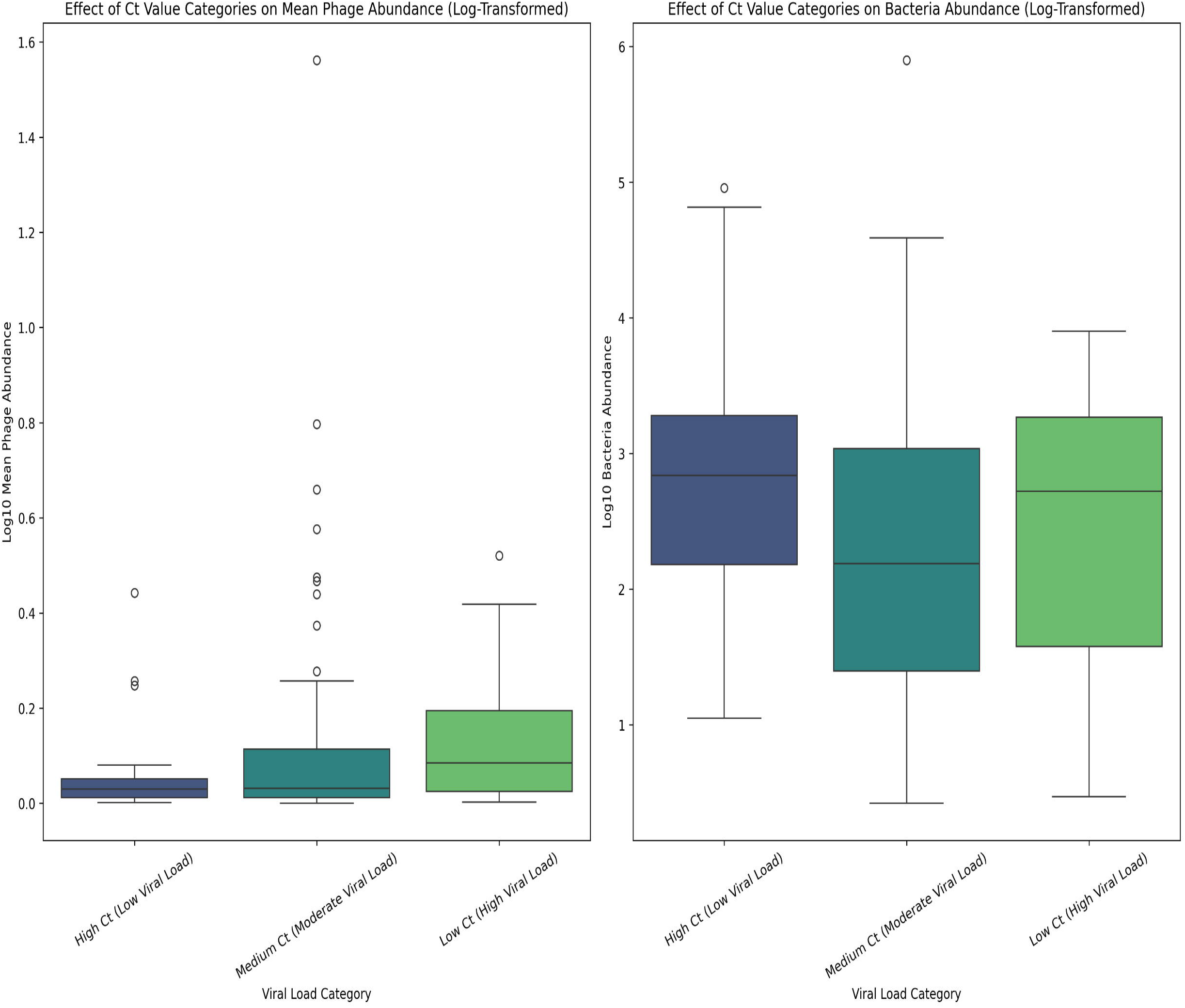
Effect of Ct Value Categories on Mean Phage and Bacteria Abundance (Log-Transformed). Boxplots showing the distribution of mean phage abundance (left) and bacterial abundance (right) across different Ct value categories. Values are log-transformed for better scaling and visualisation.

### 3.4. Disruption of Host-Phage interactions in SARS-CoV-2 group

The **(Figure 8)** represents linear and logistic regression fitting for the relationship between bacterial and bacteriophage abundance in control and SARS-CoV-2 groups. The x-axis represents the log-transformed abundance value of Bacterial host genera, and the y-axis represents the abundance of the corresponding phage genera. Panel A shows the control group, where the logistic model has an R^2^ value of 0.9265, while the linear model has an R^2^ value of 0.8906. The residual plot indicates minor deviations in the logistic fit. Panel B represents the SARS-CoV-2 group, with an R^2^ value of 0.7425 for the logistic model and 0.6440 for the linear model. The residual plots of the SARS-CoV-2 group show larger deviations in both linear and logistic models compared to the control group. The weaker fit of both linear and logistic models in the case of the SARS-CoV-2 group shows the disruption of normal host-phage interactions.

**Figure 8.**
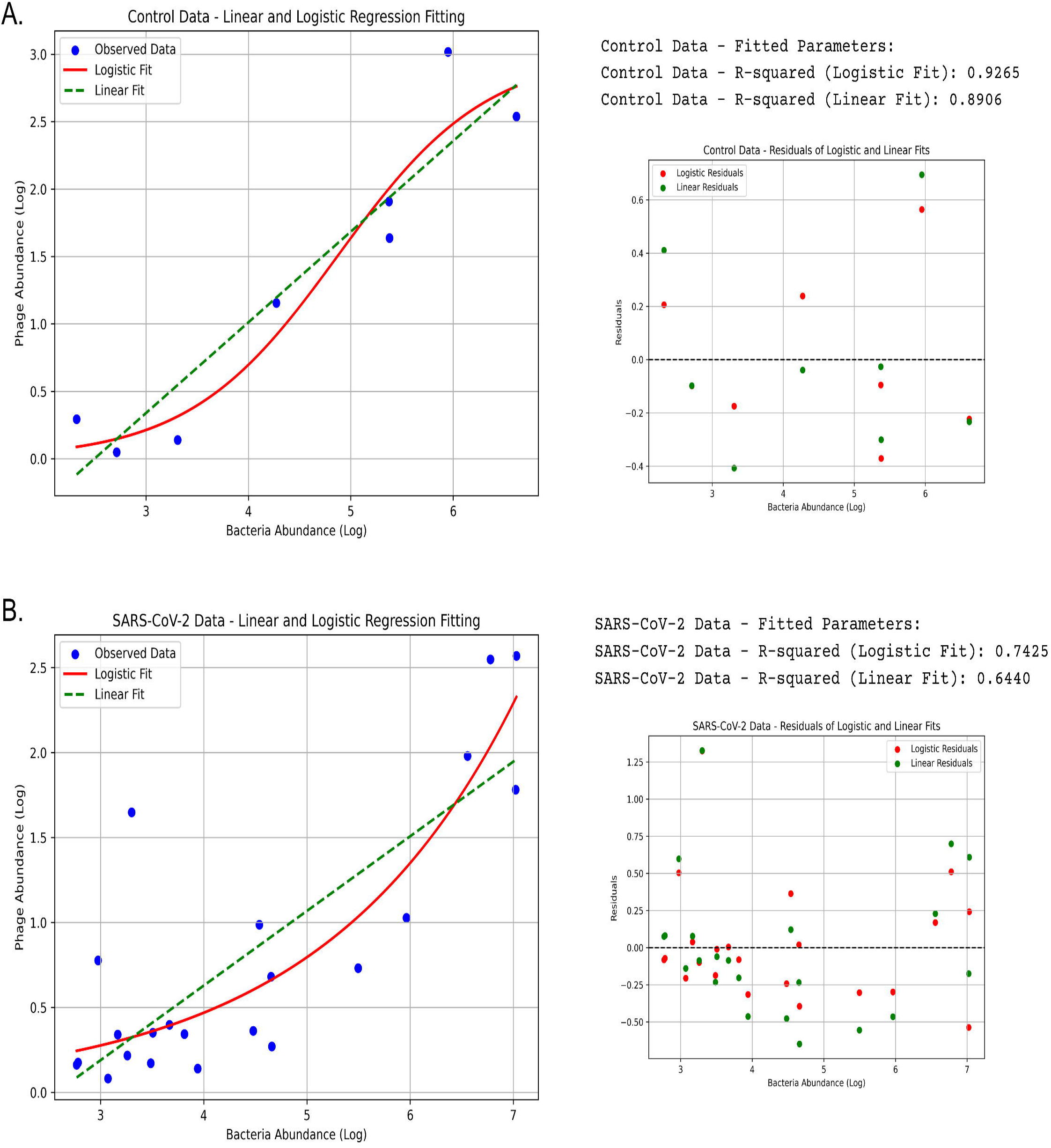
Linear and Logistic Regression Fitting for Control and SARS-CoV-2 Data. (A) Relationship between log-transformed bacteria abundance and log-transformed phage abundance in the control dataset. (B) Relationship between log-transformed bacteria abundance and log-transformed phage abundance in the SARS-CoV-2 dataset.

## 4. Discussion

Bacteriophages are one of the most ubiquitous biological entities on the earth. Bacteria and bacteriophages generally have a specific predator-prey relationship that significantly maintains the ecological balance across different microbiomes. In this study, we have tried to investigate the SARS-CoV-2-induced variations in the bacteriophage composition in the Human URT and the disruption in otherwise balanced Host-phage dynamics using a metagenomic approach. Our results have revealed significant compositional shifts among the bacterial host and phage profiles, with specific taxa being exclusive to either SARS-CoV-2 or control groups. The overall increased abundance of bacterial and phage taxa among the SARS-CoV-2 group suggests a significant shift in the microbiome, potentially driven by viral infection, immune modulation, or antibiotic use.

In our study, opportunistic bacterial genera, such as *Sphingomonas, Orchrobactrum*, and *Brevundimonas*, were exclusive to the SARS-CoV-2 group; this indicates microbiome restructuring and opportunistic colonization in response to viral infection. This finding also converges with similar studies where these bacterial taxa were consistently detected among the respiratory microbiomes of SARS-CoV-2-infected individuals [2,16,17]. The dominance of *Maxrubnervirus* in the SARS-CoV-2 group vs. *Detrevirus* in controls suggests that host bacterial changes may influence phage dynamics. The bacteriophage diversity has also increased in the SARS-CoV-2 group; higher phage richness in the SARS-CoV-2 group (79 genera vs. 28 in controls) suggests towards the adaptive response of bacteriophage community following the SARS-CoV-2 induced changes in the host diversity and increased host abundance.

c

SARS-CoV-2 samples showed higher Chao1 index values for both bacterial and phage taxa, showing greater species richness in the SARS-CoV-2 group. This increased richness could likely be a response to immune modulation or microenvironment changes in the URT of infected individuals [18,19]. The significantly lower Simpson index for bacteria in the SARS-CoV-2 group suggests that specific taxa dominate the microbiome. Meanwhile, the higher Simpson index for the SARS-COV-2 group in bacteriophage communities indicates a stable phageome. The Beta diversity analysis using the Bray Curtis dissimilarity and PCoA clearly separates the control and SARS-CoV-2 groups, showing significant microbiome variations.

The general relationship between bacteriophage and host abundance in clinical samples is not thoroughly explored; however, a recent study by Haddock et al. (2023) argued that when the abundance of pathogenic bacteria increases, the abundance of their corresponding phages also tends to increase, but not necessarily in direct proportion [20]. We have also observed a similar trend in our study, where the bacteriophage abundance tends to follow the host abundance across the samples.

The bacteriophage growth generally follows a logistic growth curve when observed over time (longitudinally); [21] However, our study attempted to ascertain the host-phage relationships among the control and SARS-CoV-2 groups in a cross-sectional context. Therefore, the linear and logistic regression models were applied to the datasets where the host and the corresponding bacteriophage abundance were X and Y variables, respectively. In the control group, the logistic regression shows a strong fit (R^2^ = 0.9265), suggesting a stable and predictable host-phage association. The linear model also exhibits a high correlation (R^2^ = 0.8906) among abundance values of bacterial hosts and their corresponding bacteriophages. Residual plots indicate minimal deviations, further supporting the structured nature of host-phage interactions. However, in the SARS-CoV-2 group, the logistic model shows weaker fit (R^2^ = 0.7425), and the linear model also shows reduced correlation (R^2^ = 0.6440), with residual plots displaying more significant deviations. This weaker correlation suggests that host-phage interactions are disrupted in SARS-CoV-2 samples, potentially due to shifts in microbial composition, altered immune responses, or environmental factors influencing viral and bacterial dynamics.

The Ct value of quantitative Reverse Transcription Polymerase Chain Reaction (qRTPCR) is a relative measure of viral load in the sample (a high Ct value corresponds to a low viral load and vice versa); However, the utility of Ct value as an indicator of disease severity is not yet fully agreed upon [22,23]. This study attempted to ascertain the relationship between the SARS-CoV-2 viral load and the abundance of bacterial hosts and corresponding bacteriophages. The Kruskal-Wallis test indicated significant differences in Mean Phage Abundance and Bacteria Abundance across Ct value categories, indicating an impact of SARS-CoV-2 viral load over Host composition. Dunn’s post-hoc analysis showed that phage abundance was significantly higher in Low Ct samples compared to Medium and High Ct groups, while bacterial abundance differed only between High and Medium Ct categories. These findings suggest that phages are more responsive to host availability, while bacterial abundance may be influenced by additional factors such as immune modulation or URT microenvironment alterations.

Identifying SARS-CoV-2-specific microbial signatures could have diagnostic or prognostic utility [24,25]. Understanding phage-bacteria interactions in the case of viral infections is essential to understanding the role of bacteriophages in complementing the immune response against the bacterial pathogens causing secondary infections. Understanding the nuances of host-phage interaction in clinical samples can help develop effective phage-based therapeutics or phage-antibiotic combinatorial therapeutic strategies for managing multidrug-resistant secondary infections during a viral pandemic [26,27]. The findings of our study highlight the need to adapt an integrative approach while developing therapeutics or preventive strategies for infectious diseases by combining virome and bacteriome data in the comprehensive context of host-microbe interaction.

## Conclusion

SARS-CoV-2 infection significantly affects the URT microbiome, altering host-phage dynamics and microbial composition. The presence of opportunistic pathogens and shifts in phage taxa suggest that viral infections could modulate host-phage interactions and the overall compositional profiles. To the best of our knowledge, this study is one of the initial studies exploring the host-phage dynamics in the upper respiratory tract using a whole genome shotgun metagenomic approach. This study emphasizes the importance of integrating virome and bacteriome analyses with potential diagnostic, prognostic, and therapeutic applications for infectious disease research.

## Limitations

Our study has a few limitations, as its cross-sectional design does not allow for assessing temporal microbiome changes. Using relative abundance metrics from the metagenomic data instead of absolute quantification limits our ability to evaluate phage-host multiplicity of infection. Other factors like immune responses, antibiotic use, etc., could influence microbiome composition, but these factors could not be controlled due to the observational and cross-sectional study design. Future research should address these gaps by integrating longitudinal studies, multi-omics approaches, and in-vitro validation of the findings.

## Data availability statement

The data is available as supplementary data with the file name “combined_supplementary_data” Any additional data, if required, will be made available on request by the authors.

## Author declaration

The authors assure that the research has followed all ethical guidelines and received approvals from the Institutional Ethics Committee for Research on Human Subjects (IEC) of CSIR-NEERI, Nagpur-20, India. Necessary consent from patients/participants has been obtained, and relevant institutional documentation has been archived. This manuscript is approved by the institutional Knowledge Resource Center (KRC) of CSIR-NEERI bearing KRC No. **CSIR-NEERI/KRC/2025/MARCH/EEPM/1**

## Confidentiality declaration

Sample IDs (23G214-5G-264_S1 to 23G214-5G-311_S48 and 24D214-5G_387_S45 to 24D214-5G_434_S91) are masked IDs and cannot be traced to participant details. The precise age of the participants is masked, and non-overlapping age ranges were used.

## Author contribution statement

SST and KK have contributed equally to the conceptualization, experimentation, and data analysis of this study.

## Conflict of interest statement

The authors declare no conflict of interest

## Acknowledgement

The authors are thankful to CSIR-NEERI for providing funds under project OLP-57 (March 2023 -April 2024) for conducting this study

## Notes

### Competing Interest Statement

The authors have declared no competing interest.

### Author Declarations

The authors assure that the research has followed all ethical guidelines and received approvals from the Institutional Ethics Committee for Research on Human Subjects (IEC) of CSIR-NEERI, Nagpur-20, India. Necessary consent from patients/participants has been obtained, and relevant institutional documentation has been archived.

